# Operational drivers of measles outbreaks in Uganda: a multi-district outbreak causality analysis, July 2025– March 2026

**DOI:** 10.64898/2026.08.06.26359852

**Authors:** Sharon Namasambi, Richard Migisha, Collins Ankunda, Yasiini Nuwamanya, Pauline Achom, Nasif Matovu, Winfred Nakaweesi, Vianney John Kigongo, Michael Mutegeki, Benon Kwesiga, Lilian Bulage, Fred Nsubuga, Brenda Nakafeero Simbwa, Immaculate Ampeire, Alex Riolexus Ario

## Abstract

**Background:** Measles outbreaks in Uganda persist despite the availability of an effective vaccine, suggesting persistent immunity gaps and health system weaknesses. We conducted a multi-district outbreak causality analysis (OCA) to identify programmatic and health-system contributors to measles outbreaks and inform measles elimination programming.

**Methods:** We conducted a cross-sectional mixed-methods OCA across 12 affected districts in Uganda (2025–2026), guided by the World Health Organization framework. We reviewed measles case investigation reports and triangulated findings with qualitative interviews with district health teams, health workers, surveillance and immunisation staff, community leaders, Village Health Teams, and caregivers. We deductively analysed data to identify causal pathways and contributing factors. Findings were organized into four prespecified analytical themes: immunization service delivery, caregiver access and demand, surveillance and case detection, and outbreak preparedness and response.

**Results:** The 12 districts reported 1,302 cases, including 80 laboratory-confirmed cases and 10 suspected deaths (case-fatality rate: 0.77%); 46.5% (n=606) occurred among children aged 18– 59 months. Most cases (65.3%) occurred in unvaccinated children, versus 1.0% in children with both measles-rubella (MR) doses. Across districts, incomplete MR2 implementation, irregular outreach, inconsistent fixed-site vaccination, weak follow-up of children who missed vaccination, and transport and distance barriers contributed to persistent immunity gaps. Low clinical suspicion, limited engagement of Village Health Teams, laboratory and surveillance-information bottlenecks, absence of pre-positioned response plans, delayed response activation, and inadequate isolation capacity further limited early detection and control.

**Conclusion:** Measles outbreaks were driven primarily by missed vaccination, especially incomplete delivery of the two-dose MR schedule, compounded by access barriers, delayed case detection, and limited outbreak preparedness. Strengthening routine MR1 and MR2 delivery, targeted catch-up vaccination, community-linked surveillance, and pre-positioned district response plans with clear activation triggers will be critical to closing immunity gaps and accelerating measles elimination in Uganda.

## Introduction

Measles is a highly contagious viral disease and continues to cause substantial morbidity and mortality despite the availability of a safe and highly effective vaccine (1,2). Sustained interruption of measles transmission requires high population immunity, generally achieved through at least 95% of two doses of measles containing vaccine delivered through routine immunisation and, when needed, supplementary immunization activities (3,4). However, recent years have witnessed a global resurgence of measles. In 2024, the World Health Organization (WHO) reported approximately 395,000 laboratory confirmed cases worldwide (4), reflecting the growing accumulation of susceptible populations following disruptions to routine immunisation, surveillance challenges, and inequitable access to vaccination services (4–6). The burden has been particularly pronounced in sub-Saharan African countries that recorded a substantial proportion of reported cases during early 2025 (7). These recurrent outbreaks may largely reflect accumulated immunity gaps driven by complex programmatic, health system, and contextual factors, emphasising the need to move beyond descriptive outbreak investigations toward systematic identification of their underlying causes (8).

Uganda has achieved considerable progress in expanding immunization services; however, routine measles vaccination coverage remains insufficient to achieve elimination targets. National estimates indicate that coverage for the first dose of the measles rubella vaccine (MR1) reached 93% in 2023, whereas coverage for the second dose (MR2) remained substantially lower at 21% in 2024, increasing to approximately 54% by March 2025 (9). These persistent immunity gaps have been accompanied by recurrent outbreaks across multiple districts. By the end of 2025, confirmed measles outbreaks had been reported in 66 districts, with transmission interrupted in only 37 districts at the time of analysis. Previous investigations in Uganda have identified immediate determinants of transmission, including non-vaccination, household exposure, health facility transmission, and congregation of susceptible children (10–13).

Although conventional outbreak investigations provide essential information on the magnitude, distribution, and immediate risk factors of outbreaks, they often do not identify the structural and programmatic conditions that allow outbreaks to emerge and persist. To address this limitation, the WHO Measles Outbreak Guide recommends structured outbreak causality analysis to systematically identify deficiencies in immunisation services, surveillance systems, outbreak response, governance, and community engagement that contribute to recurrent transmission (8). This approach is particularly useful where outbreak investigations repeatedly identify missed vaccination but do not sufficiently explain why children remain unvaccinated or incompletely vaccinated. We conducted a multi-district outbreak causality analysis of measles outbreaks reported in selected Ugandan districts during July 2025–March 2026, to identify operational and health-system contributors to measles transmission, and generate recommendations for strengthening measles elimination programming in Uganda.

## Methods

### Study design

We conducted a cross-sectional mixed-methods study nested within routine measles outbreak investigations conducted between July 2025 and March 2026. This period coincided with the introduction of the WHO Outbreak Causality Analysis (OCA) framework into Uganda’s measles outbreak response, providing a standardised approach to identify underlying programmatic, health system, and community-level factors contributing to measles transmission beyond routine epidemiologic assessment (8).

### Study setting and district inclusion

The study was conducted in 12 Ugandan districts with confirmed ongoing measles outbreaks during the study period (Figure 1). These were all districts in which the WHO OCA framework was incorporated into routine measles outbreak investigations between July 2025 and March 2026. District inclusion was therefore based on two operational criteria: confirmation of an ongoing measles outbreak and implementation of an OCA. Districts were not selected according to outbreak size, recurrence, or geographic location.

**Figure 1:**
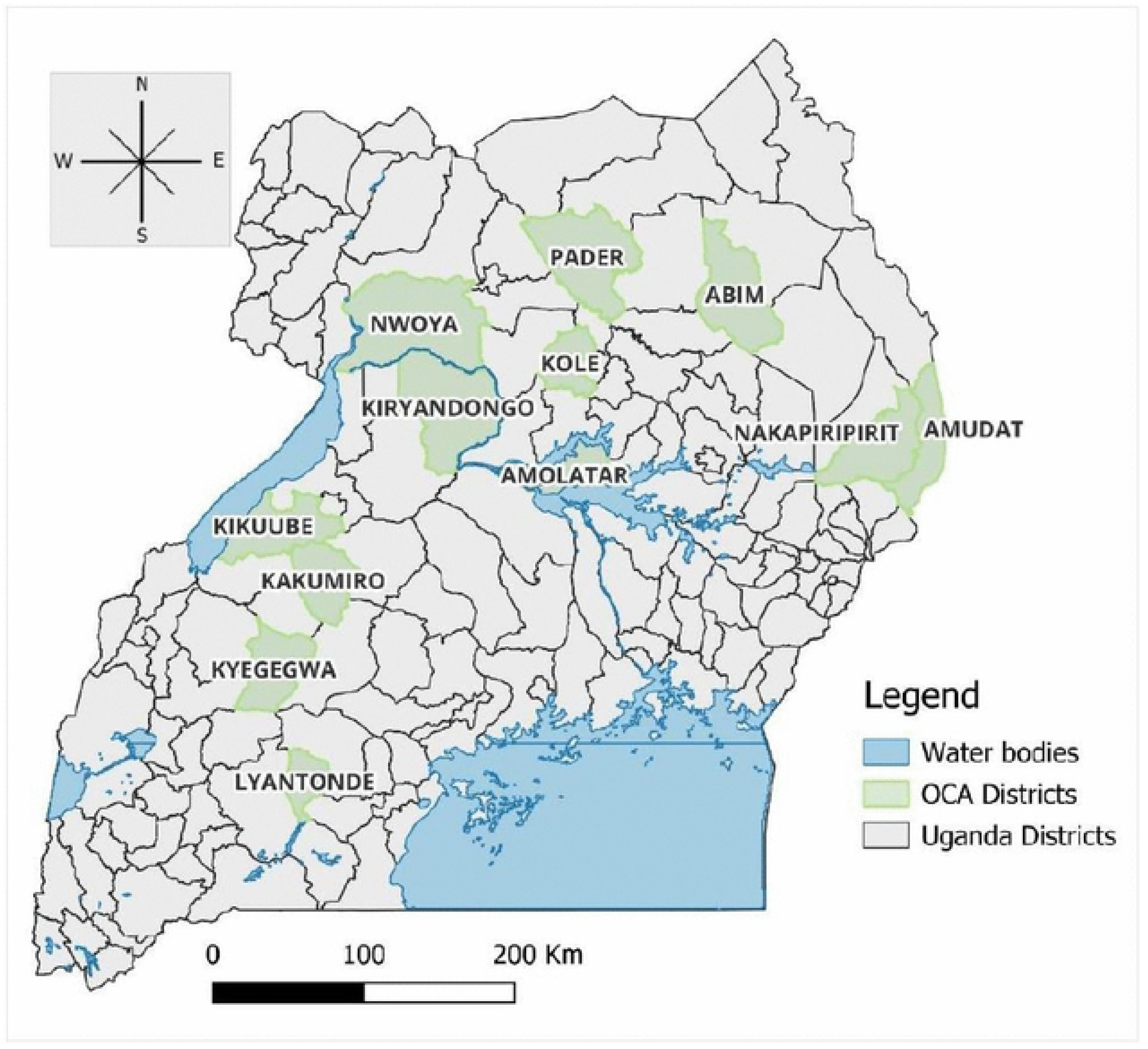
Map of Uganda showing the 12 districts assessed for outbreak causality analysis (OCA), July 2025– March 2026.

### Case and outbreak definitions

A suspected measles case was defined according to Uganda’s national Integrated Disease Surveillance and Response guidelines as illness in a resident of an assessed district during the outbreak period characterised by fever and a generalised maculopapular rash, with at least one of the following: cough, coryza, or conjunctivitis (8,14).

A laboratory-confirmed measles case was defined as a suspected case with detection of measles-specific IgM antibodies from serum by laboratory testing.

A suspected measles-associated death was defined as death occurring in a person meeting the suspected measles case definition, where no alternative confirmed cause of death was established.

A measles outbreak was considered confirmed when a district reported three or more laboratory-confirmed measles cases within a 30-day period in the same district or epidemiologically linked sub-district, consistent with Uganda’s national measles outbreak investigation and response criteria. Outbreak confirmation was based on routine surveillance information generated through district surveillance systems, verified through laboratory confirmation by the Uganda Virus Research Institute (UVRI) measles laboratory, and formally recognised through the Ministry of Health (MoH) surveillance and outbreak declaration mechanisms.

### Study population

The study population comprised key stakeholders involved in measles prevention and outbreak response, including members of the District Health Team (DHT), health facility staff, community leaders and gatekeepers, and caregivers of measles cases.

### Sample size and sampling methods

Participants were purposively selected based on their roles in measles surveillance, outbreak detection and response, immunisation service delivery, or community engagement. In each of the 12 outbreak districts, one focus group discussion (FGD) comprising 6–10 district health team members and health facility staff was conducted. Key informant interviews (KIIs) were conducted with surveillance focal persons, immunisation officers, and district leaders selected for their technical expertise and decision-making roles. At the community level, one FGD involving 5–6 caregivers of children with measles explored care-seeking and immunisation experiences, while KIIs with community gatekeepers, including local leaders, Village Health Team members, and religious leaders, provided contextual perspectives on factors contributing to the outbreaks. Approximately five KIIs were conducted per district, yielding 60 KIIs in total across the 12 districts.

### Root cause analysis framework

The analysis was guided by the WHO Measles Outbreak Causality Analysis (OCA) framework (8). The framework assesses measles outbreaks through a two-level approach. At Level 1, outbreaks are evaluated for three primary determinants: surveillance gaps, outbreak response gaps, and immunity gaps. Where immunity gaps are identified, Level 2 analysis examines whether these result from failure to vaccinate or vaccination failure and characterises underlying causes according to policy, provider, and client-level factors (Figure 2).

**Figure 2:**
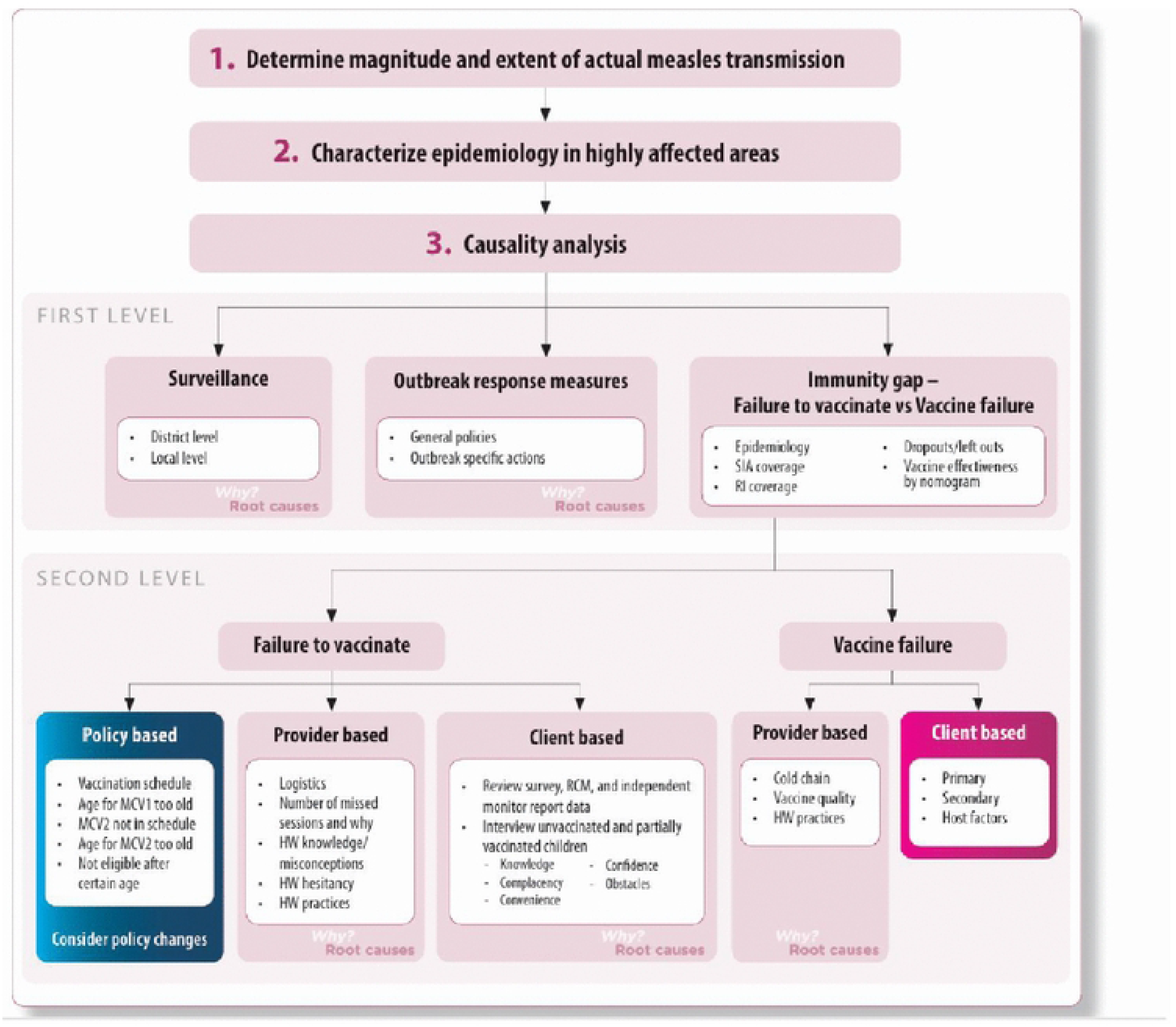
Outbreak Causality Analysis (OCA) Framework, adopted from WHO, 2022.

### Data collection

Qualitative data were collected through key informant interviews (KIIs) and focus group discussions (FGDs) using semi-structured guides adapted from the WHO Measles Outbreak Causality Analysis (OCA) framework. The guides explored determinants of measles outbreaks across surveillance, outbreak response, and immunity gap domains. Interviews and FGDs were conducted in English by trained multidisciplinary outbreak investigation teams comprising epidemiologists and surveillance officers. Participants were purposively selected based on their roles in outbreak detection, immunization service delivery, and community engagement.

Discussions were audio-recorded and documented using detailed field notes. Qualitative findings were triangulated with routine immunization and surveillance data, including health facility registers, DHIS2/HMIS immunization coverage reports, measles case investigation forms, and laboratory records.

### Health facility assessment

Health facility assessments were conducted using the WHO OCA facility assessment checklist to evaluate potential programmatic causes of vaccine failure. Assessments included review of cold-chain equipment functionality, temperature monitoring records, vaccine storage and handling practices, vaccine stock availability, immunization session organization, and vaccination practices through direct observation and interviews with vaccination staff.

### Data management and analysis

Quantitative data from case line lists, case investigation forms, laboratory records, and routine immunisation reports were checked for completeness and consistency. We summarised case characteristics using frequencies and percentages. Vaccination status was categorised as no recorded MR dose, one recorded dose, two recorded doses, or unknown or undocumented status. The suspected case-fatality proportion was calculated by dividing the number of suspected measles-associated deaths by the total number of reported measles cases. District MR1 and MR2 administrative coverage estimates were summarised using ranges and compared with the ≥95% coverage target required for measles elimination.

Root causes identified from individual district assessments were synthesized across districts using the multi-district approach described in the WHO Measles Outbreak Guide (2022) (8). Similar contributing factors were harmonized across districts and summarized according to their frequency of occurrence to identify common operational drivers of measles outbreaks. Qualitative data from KIIs and FGDs were analysed using a deductive thematic approach guided by the WHO Measles Outbreak Causality Analysis (OCA) framework. For this analysis, the OCA framework was operationalised into four prespecified analytical themes including; immunization service delivery, caregiver access and demand, surveillance and case detection, and outbreak preparedness and response to guide data abstraction, interpretation, and presentation of findings. Immunization service delivery and caregiver access and demand themes captured Level 2 determinants contributing to failure to vaccinate, while surveillance and outbreak preparedness and response corresponded to Level 1 outbreak system gaps.

A coding framework based on these themes was applied to transcripts and field notes, while allowing refinement of subthemes where supported by the data. Data collection and analysis continued until thematic saturation was reached, with no new major themes emerging from subsequent district assessments.

## Results

### Outbreak profile and vaccination status

The outbreak causality analyses were conducted in 12 districts across six regions of Uganda. These districts reported 1,302 measles cases, including 80 laboratory-confirmed cases and 10 suspected measles-associated deaths. Case counts ranged from 41 in Nakapiripirit to 245 in Lyantonde, with the highest burdens observed in Lyantonde (n=245) and Kole (n=185) Districts. Children aged 18–59 months accounted for nearly half of all reported cases (n=606; 46.5%). Substantial immunity gaps were evident among cases: 850 (65.3%) had received no MR vaccine dose, while only 13 (1.0%) had completed the recommended two-dose MR schedule. Routine MR coverage remained below the 95% threshold required to interrupt measles transmission in all 12 districts (Table 1).

**Table 1.** Epidemiologic profile of measles outbreaks in 12 study districts, Uganda, July 2025–March 2026 (N=1,302)

### Cross-district operational contributors

Cross-district synthesis identified recurrent operational contributors across the four prespecified analytical themes (Table 2). Seven contributors were identified in all 12 districts, including inadequate MR2 integration into routine immunization services, irregular outreach vaccination, transport and distance barriers to vaccination, low clinician suspicion for measles, weak Village Health Team (VHT) engagement in surveillance, absence of pre-positioned response plans, and delayed outbreak response activation. Additional contributors identified in most districts (10–11/12) included inconsistent daily static immunization services, inadequate vaccination microplanning, weak MR1-to-MR2 defaulter tracking, and poor infection prevention and isolation practices. In contrast, vaccine-related misconceptions were identified in six districts, while lack of a vaccinating health facility was reported in only three districts (Figure 3).

**Table 2:** Cross-district operational contributors to measles outbreaks across 12 assessed districts, Uganda, July 2025-March 2026.

**Figure 3:**
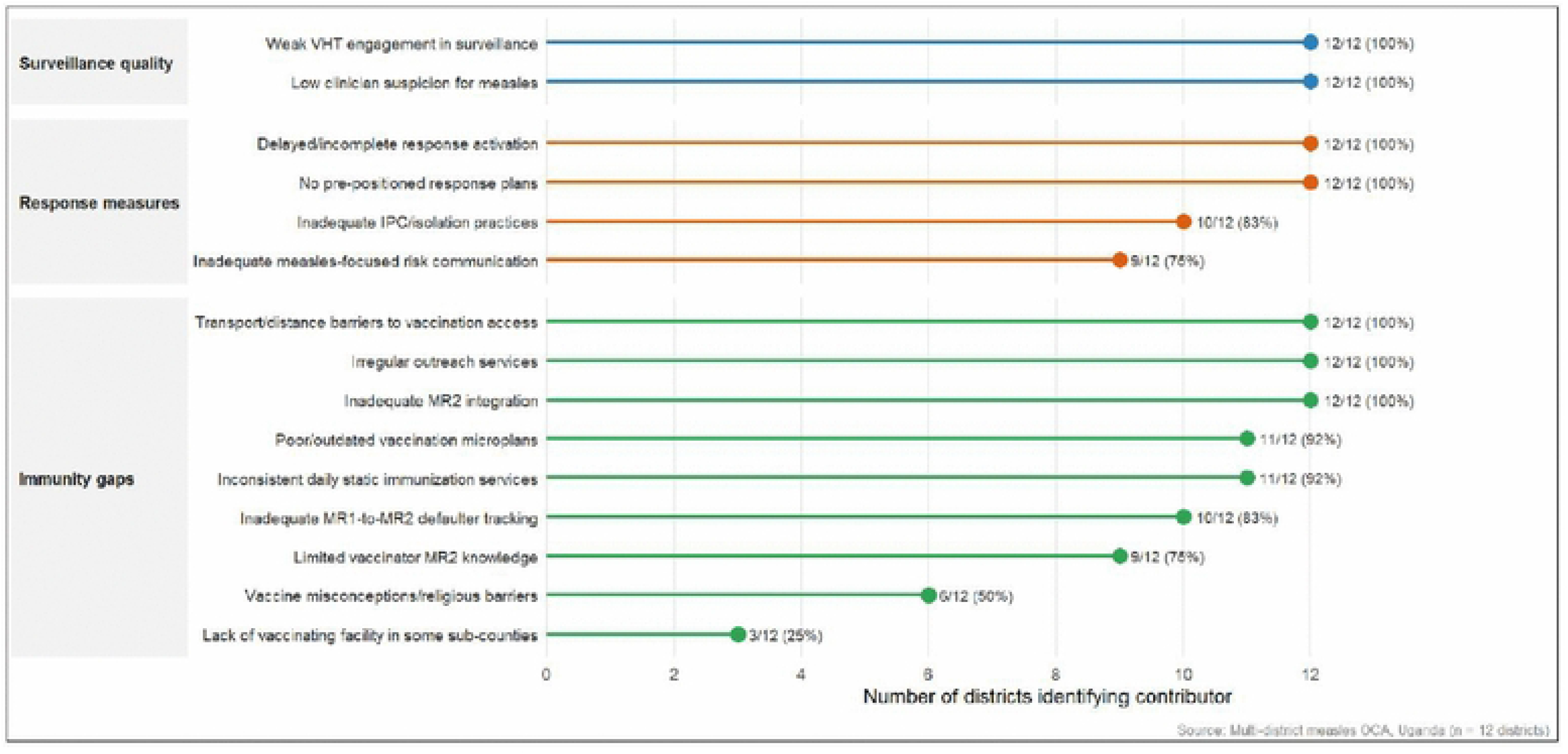
Cross-district frequency of identified operational contributors to measles outbreaks across 12 assessed districts, Uganda, July 2025-March 2026.

### Immunisation service delivery gaps

Immunization service delivery gaps were the predominant supply-side contributors to immunity gaps. Although MR2 had been incorporated into the national immunisation schedule, implementation remained inconsistent because vaccinators had limited awareness of the updated schedule, dissemination of revised guidance was inadequate, supportive supervision was limited, and systems to identify children who missed MR2 were weak

> *“Vaccination is mainly left to the vaccinators, who are usually CHEWs, VHTs, or Nursing Assistants. Many of them are not fully updated on the new schedules, including MR2, and supervision from the technical team is limited.” FGD 11, P4*

Routine vaccination delivery was further weakened by irregular outreach services, inconsistent daily static immunisation, weak or outdated microplanning, and last-mile supply constraints. Outreach was intended to reach underserved communities but was often delayed because of transport, fuel, and operational funding constraints. In some facilities, vaccination was offered on selected days only, creating missed opportunities when caregivers brought children for other services on non-immunisation days.

> *“Outreach services are supposed to help the hard-to-reach areas, but they are not conducted regularly because transport and facilitation are limited.” FGD 12, P4*
>
> *“Sometimes immunisation is done on specific days instead of daily, so if a caregiver comes on another day, the child may be asked to return later.” KII 4, P1*

Health facility assessments found no major evidence of programmatic vaccine failure.

Cold-chain equipment was functional, vaccine vial monitors were within acceptable limits, and observed vaccine storage, reconstitution, and administration practices were consistent with national guidelines.

### Caregiver access and demand barriers

Caregiver-related contributors were predominantly structural rather than behavioral. Across all districts, transport costs, long distances to vaccination points, dependence on outreach services, and the opportunity costs of repeated facility visits limited access to vaccination, even among caregivers willing to vaccinate their children.

> *“We want to vaccinate our children, but the distance to the facility makes it difficult, especially when there is no money for transport.” FGD 10, CG3*

Limited awareness of the two-dose MR schedule further contributed to incomplete vaccination. Caregivers frequently reported knowing about MR1 but not the need for a second dose because counselling on return dates and community reminder systems were inadequate as noted by participants.

> *“We only knew about the first measles vaccine. We did not know the child is supposed to return for another dose.” FGD 5, CG1*

Vaccine-related misconceptions associated with cultural and religious beliefs were reported in six districts but were less widespread than access barriers and weaknesses in routine service delivery.

> *“Measles is seen as a traditional illness caused by spiritual factors, so some caregivers prefer traditional remedies and are concerned that vaccines may harm the child.” FGD 12, CG4*

### Surveillance and case detection gaps

Delayed case detection resulted from weaknesses at multiple levels of the surveillance system. At health facilities, clinicians frequently reported low suspicion for measles, limited refresher training on case definitions, and initial misclassification of febrile rash illnesses. One of the participants noted;

> *“When the first children came with fever and rash, measles was not the first thing that came to mind. Many of us thought it was other skin conditions or a common viral infection, so the suspicion for measles came quite late.” FGD 10, P2*

Community-based surveillance was similarly weak. VHTs reported limited orientation on measles case definitions, inadequate facilitation for active case finding, and weak communication with health facilities and district surveillance teams.

> *“Because we were not trained on the measles case definition, we only reported a few children when the outbreak was already spreading.” KII 12, P1*

Delayed specimen collection, prolonged laboratory turnaround times, and limited access to surveillance information systems further delayed outbreak confirmation and response.

> *“Some of the samples we sent never came back with results. Because of this, the team became less alert and did not actively search for more suspected cases.” KII 11, P3*

### Outbreak preparedness and response gaps

Preparedness for measles outbreaks was limited across all assessed districts. Most districts lacked pre-positioned response plans, clearly defined activation triggers, and district-specific outbreak microplans, resulting in delayed initiation of reactive vaccination and other control measures.

> *“By the time the response planning started, the outbreak had already spread widely. We did not have a ready microplan to guide the vaccination response.” FGD 9, P1*

Response implementation was further constrained by weak coordination and accountability. Task force decisions were not consistently communicated to operational teams, key logistics and cold-chain personnel were engaged late, and agreed actions were not uniformly implemented across affected areas.

> *“Sometimes decisions are made during the task force meetings, but the information does not reach the facilities in time for action.” KII 12, P5*
>
> *“The cold chain team was not aware that an outbreak had already been declared, so vaccine preparation for the response did not start immediately.” KII 11, P2*

Infection prevention and control capacity was also inadequate. Suspected measles cases were frequently managed alongside other pediatric patients because of limited isolation space and delayed triage, while some households received little guidance on home isolation.

> *“Because space is limited in the paediatric ward, it becomes difficult to separate suspected measles cases from other patients.” KII 6, P1*
>
> *“Some of the children were infected while already admitted on the ward before an isolation area was identified.” KII 9, P3*

## Discussion

Measles outbreaks across the 12 Ugandan districts were mainly driven by gaps in programme delivery rather than vaccine availability. Most cases occurred among children with no recorded MR vaccination, while completion of the recommended two-dose schedule was rare. Inconsistent MR2 delivery, unreliable outreach, weak follow-up of children who missed vaccination, transport barriers, delayed clinical recognition, limited community surveillance, and inadequate response readiness likely interacted to sustain transmission. These findings highlight modifiable weaknesses across routine immunisation, surveillance, and district preparedness.

The high proportion of zero-dose and under-vaccinated cases supports failure to vaccinate, rather than vaccine failure, as the principal source of persistent measles susceptibility in Uganda. This is in agreement with regional evidence showing that measles outbreaks persist where two-dose coverage remains insufficient to maintain elimination-level population immunity, despite the high effectiveness of measles vaccines (4,11,15–17). In the present analysis, missed opportunities and inconsistent service availability appeared more prominent than widespread hesitancy. Closing these gaps will require equitable access to dependable routine and outreach services, particularly for communities facing transport and distance barriers (18,19).

Despite formal inclusion of MR2 in Uganda’s national immunisation schedule, operationalisation remained incomplete. Weak microplanning, limited caregiver awareness, inadequate supportive supervision, and poor follow-up of children who missed vaccination constrained routine delivery. This is consistent with experience elsewhere in Africa, where policy introduction has often preceded the development of sufficient delivery and monitoring capacity (20,21). Effective implementation will require sustained investment in workforce capacity, supervision, microplanning, and systems that track and support completion of the two-dose schedule.

Community-level barriers were predominantly structural. Distance, transport costs, and inconsistent outreach reduced access even among caregivers willing to vaccinate, consistent with evidence that inequitable service access is a major determinant of under-immunisation in many low-income settings (19,22,23). Hesitancy linked to cultural or religious beliefs was identified in some communities but was less consistently reported. Coverage improvements will therefore depend primarily on expanding reliable outreach, locating services closer to underserved populations, and strengthening follow-up of children who miss vaccination.

Surveillance weaknesses contributed substantially to delayed outbreak detection. Low clinical suspicion, inconsistent application of surveillance guidelines, and limited VHT engagement reduced opportunities to identify transmission early, consistent with evidence from other resource-limited settings (8,10,11,17). Strengthening surveillance will require more than improved reporting platforms; it will also depend on continuous development of frontline clinical capacity (24–27), sustained support for community-based surveillance, and timely access to laboratory and surveillance information to guide rapid response.

Weak preparedness reduced the timeliness and effectiveness of outbreak control. The absence of pre-positioned response plans, predefined activation triggers, and clearly assigned responsibilities meant that planning frequently began only after transmission was well established, indicating that preparedness had not been adequately institutionalised within routine district health systems. This is consistent with evidence that advance preparedness is associated with faster containment (15,28). Delayed recognition of suspected cases, coupled with inadequate isolation practices, created opportunities for healthcare-associated transmission, mirroring nosocomial amplification documented in previous Uganda outbreaks and other resource-constrained settings (13,29,30). Embedding preparedness within routine district emergency planning, with regular simulation exercises and reviews, and integrating infection prevention and control, rapid triage, and isolation protocols into outbreak preparedness, could improve response timeliness and reduce facility-associated spread.

Taken together, these findings suggest that recurrent measles outbreaks in Uganda are not isolated epidemiological events but manifestations of persistent weaknesses across the immunisation and public health system. Achieving measles elimination will require coordinated investments that strengthen routine immunisation, particularly effective MR2 implementation, improve surveillance sensitivity, institutionalise outbreak preparedness, and ensure equitable access to vaccination services.

## Limitations

This analysis has some limitations. The 12 districts represented all settings in which the OCA framework was implemented during the study period, but they may not represent all measles-affected districts in Uganda, particularly those where outbreaks were not detected, investigated, or assessed using the framework. Vaccination status and district coverage estimates were derived partly from routine records and caregiver reports, which were incomplete for some case-patients and may have introduced recall, reporting, or misclassification bias. The cross-sectional mixed-methods design identified recurring and plausible operational pathways but could not establish causality or quantify the independent contribution of each factor to outbreak occurrence or duration. In addition, host factors that may influence vaccine protection, including nutritional status, HIV infection, and other causes of immune impairment, were not systematically assessed; therefore, vaccine failure could not be completely excluded. Nevertheless, use of a standardised framework and triangulation across surveillance and immunisation records, laboratory data, interviews, focus group discussions, and health-facility assessments strengthened the consistency, credibility, and programmatic relevance of the findings.

## Conclusion

Measles outbreaks across the assessed districts reflected preventable gaps in vaccination, surveillance, and preparedness. Inadequate delivery of MR1 and MR2, missed opportunities for vaccination, weak surveillance, and limited response readiness left susceptible children unprotected and allowed transmission to continue. Progress towards elimination will require sustained routine immunisation, targeted recovery of zero-dose and under-vaccinated children, timely community-linked detection, and institutionalised district preparedness. Future studies could evaluate the effectiveness, feasibility, and cost of interventions designed to improve MR2 uptake, reach missed children, strengthen early detection, and shorten the time from outbreak recognition to response.

## Declarations

### Ethics approval and accordance

This causality analysis was conducted as part of a public health emergency response and was approved as public health practice by the Uganda Ministry of Health and determined to be non-research by the Centers for Disease Control and Prevention Human Subjects Review Board (National Institute for Occupational Safety and Health Institutional Review Board). The study was conducted in accordance with applicable federal laws and CDC policy (45 C.F.R. part 46; 21 C.F.R. part 56; 42 U.S.C. §241(d)). All human-related procedures were carried out in accordance with the Declaration of Helsinki and the Council for International Organizations of Medical Sciences (CIOMS) International Ethical Guidelines.

### Consent to participate

Informed consent was obtained from all participants aged ≥18 years.

### Consent for publication

Not applicable

### Clinical Trial Number

Not applicable

### Data availability

The datasets generated and analysed during this study are the property of the Uganda Public Health Fellowship Program and are not publicly available to protect participant confidentiality.

Data can be made available from the corresponding author upon reasonable request, subject to approval by the Uganda Public Health Fellowship Program.

### Author contributions

SN conceptualized the study, led data collection and analysis, and drafted the manuscript. PA, WN, MM, NM and VJK supported data collection, analysis, and report development. RM, CA, and YN provided scientific review of the initial field report and the manuscript draft. LB, RM, BK, YN, FN, IA, and ARA contributed to manuscript review and editing. All authors read and approved the final manuscript.

## Acknowledgements

We acknowledge the Uganda Expanded Program on Immunization (UNEPI), Ministry of Health, the Global Alliance for Vaccines and Immunization (GAVI), and the Uganda Public Health Fellowship Program for their technical guidance. We also thank all District Health Teams for their commitment and support during this investigation.

## Disclaimer

The findings and conclusions presented in this report are those of the authors and do not necessarily reflect the official views of the US Centers for Disease Control and Prevention or the Uganda Ministry of Health.

## Abbreviations

DHT: District Health Team
FGD: Focus Group Discussion
MR: Measles-Rubella vaccine
MoH: Ministry of Health
US: United States
CDC: Centers for Disease Control and Prevention
OCA: Outbreak Causality Analysis
RCA: Root Cause Analysis
VHT: Village Health Team
KII: Key Informant Interview
WHO: World Health Organization
MCV: Measles Containing Vaccine

## References

1. CDC. Measles (Rubeola) [Internet]. 2025 [cited 2025 Jul 21]. Measles Cases and Outbreaks. Available from: https://www.cdc.gov/measles/data-research/index.html

2. Guerra FM, Bolotin S, Lim G, Heffernan J, Deeks SL, Li Y, et al. The basic reproduction number (R0) of measles: a systematic review. Lancet Infect Dis. 2017 Dec 1;17(12):e420–8. doi:10.1016/S1473-3099(17)30307-9 PubMed PMID: 28757186.

3. Cutts FT, Ferrari MJ, Krause LK, Tatem AJ, Mosser JF. Vaccination strategies for measles control and elimination: time to strengthen local initiatives. BMC Med. 2021 Jan 5;19(1):2. doi:10.1186/s12916-020-01843-z

4. Do LAH, Mulholland K. Measles 2025. N Engl J Med. 0(0). doi:10.1056/NEJMra2504516

5. World Health Organisation. What Does it Take to Track and Tame Measles Outbreaks? – WHO Foundation [Internet]. 2026 [cited 2026 Mar 12]. Available from: https://www.who.foundation/post/what-does-it-take-to-track-and-tame-measles-outbreaks

6. UNICEF and WHO warn of perfect storm of conditions for measles outbreaks, affecting children [Internet]. [cited 2025 Nov 8]. Available from: https://www.who.int/news/item/27-04-2022-unicef-and-who-warn-of--perfect-storm--of-conditions-for-measles-outbreaks--affecting-children

7. World Health Organization AFRO. Measles AFRO Epidemiological Update [Internet]. 2025. Available from: https://dataportal.afro.who.int/wp-content/uploads/2025/04/Measles_Report.pdf

8. World Health Organisation. Measles Outbreak Guide [Internet]. 1st ed. Geneva: World Health Organization; 2022. 1 p. Available from: https://iris.who.int/server/api/core/bitstreams/d37add05-b90c-4f37-a115-e35514e95963/content

9. World Health Organization, United Nations Children’s Fund (UNICEF). Progress and Challenges with Achieving Universal Immunization Coverage: WHO/UNICEF Estimates of National Immunization Coverage (WUENIC), 2023. Geneva: WHO; 2024. [Internet]. WHO/UNICEF; 2024. Available from: https://cdn.who.int/media/docs/default-source/immunization/wuenic/wuenic-progress-and-challenges.pdf?sfvrsn=d6907baa_9&download=true

10. Okiror EO, Ampaire I, Nsubuga F, Kwizera P, Okello PE, Migisha R, et al. Measles outbreak with children below the recommended age for first dose of measles vaccine most affected in Kakumiro District, February–May 2024. Vol. 9. 2024;9(3).

11. Mfitundinda E, Migisha R, Namusisi AM, Wenani D, Kwesiga B, Ayeerwot R, et al. Investigation of a cross-border measles outbreak in Moroto District, northeastern Uganda, March–September, 2024. Discov Public Health. 2025 May 16;22(1):267. doi:10.1186/s12982-025-00581-y

12. Namasambi S, Migisha R, Kigongo VJ, Nakabuye M, Bulage L, Kwesiga B, et al. School-associated measles outbreak driven by vaccination gaps, sociocultural barriers, and delayed detection in Gomba District, Uganda 2025. Discov Public Health. 2026 Jul 10;23(1):1168. doi:10.1186/s12982-026-02544-3

13. Namusisi AM, Nuwamanya Y, Migisha R, Nsubuga F, Baganizi M, Ampeire I, et al. Measles outbreak investigation in Terego District, Uganda, February ─June, 2024. Discov Public Health. 2026 May 22;23(1):747. doi:10.1186/s12982-026-02085-9

14. Ministry of Health. Uganda 3rd IDSR Tech Guideline_PrintVersion_10Sep2021.pd [Document] [Internet]. Ministry of Health; 2021. Available from: https://www.afro.who.int/sites/default/files/2021-09/2_Uganda%203rd%20IDSR%20Tech%20Guideline_PrintVersion_10Sep2021.pdf

15. Minta AA, Ferrari M, Antoni S, Lambert B, Sayi TS, Hsu CH, et al. Progress Toward Measles Elimination — Worldwide, 2000–2023. Vol. 73. 2024;73(45).

16. Minta AA. Progress Toward Measles Elimination — Worldwide, 2000–2022. MMWR Morb Mortal Wkly Rep. 2023;72. doi:10.15585/mmwr.mm7246a3

17. Kabami Z, Simbwa BN, Kizito SN, Agaba B, Kayiwa J, Kadobera D, et al. Epidemiological characteristics and trends of measles cases reported through the case-based surveillance system, Uganda, 2016 – 2020. Vol. 8. 2023;8(4).

18. Adisu MA. Timeliness of the second dose of measles-containing vaccine uptake and its determinants among children aged 24–36 months in Gondar City, Northwest Ethiopia, 2023: Community-based cross-sectional study design. J Virus Erad. 2025 Jun 1;11(2):100594. doi:10.1016/j.jve.2025.100594

19. Alemu TG, Tamir TT, Workneh BS, Mekonen EG, Ali MS, Zegeye AF, et al. Coverage and determinants of second-dose measles vaccination among under-five children in East Africa countries: a systematic review and meta-analysis. Front Public Health. 2024 May 1;12:1359572. doi:10.3389/fpubh.2024.1359572 PubMed PMID: 38751581; PubMed Central PMCID: PMC11094336.

20. Masresha BG, Shibeshi ME, Grant GB, Hatcher C, Wiysonge CS. Progress with the Second Dose Measles Vaccine Introduction and Coverage in the WHO African Region. Vaccines. 2024 Sep 18;12(9):1069. doi:10.3390/vaccines12091069 PubMed PMID: 39340099; PubMed Central PMCID: PMC11435470.

21. Portnoy A, Jit M, Helleringer S, Verguet S. Impact of measles supplementary immunization activities on reaching children missed by routine programs. Vaccine. 2018 Jan 2;36(1):170–8. doi:10.1016/j.vaccine.2017.10.080 PubMed PMID: 29174680; PubMed Central PMCID: PMC5949217.

22. Ozawa S, Yemeke TT, Evans DR, Pallas SE, Wallace AS, Lee BY. Defining hard-to-reach populations for vaccination. Vaccine. 2019;37(37):5525–34.

23. Larson HJ, Jarrett C, Eckersberger E, Smith DMD, Paterson P. Understanding vaccine hesitancy around vaccines and vaccination from a global perspective: A systematic review of published literature, 2007–2012. Vaccine. 2014 Apr 17;32(19):2150–9. doi:10.1016/j.vaccine.2014.01.081

24. says M. Uganda Village Health Teams Programme. Uganda’s Village Health Teams Program. CHW Central [Internet]. 2021 [cited 2026 Mar 18]. Available from: https://chwcentral.org/ugandas-village-health-teams-program/

25. Mays DC, O’Neil EJ, Mworozi EA, Lough BJ, Tabb ZJ, Whitlock AE, et al. Supporting and retaining Village Health Teams: an assessment of a community health worker program in two Ugandan districts. Int J Equity Health. 2017 Jul 20;16:129. doi:10.1186/s12939-017-0619-6 PubMed PMID: 28728553; PubMed Central PMCID: PMC5520299.

26. Pandya S, Hamal M, Abuya T, Kintu R, Mwanga D, Warren CE, et al. Understanding Factors That Support Community Health Worker Motivation, Job Satisfaction, and Performance in Three Ugandan Districts: Opportunities for Strengthening Uganda’s Community Health Worker Program. Int J Health Policy Manag. 2022 Apr 18;1. doi:10.34172/ijhpm.2022.6219

27. Masiira B, Nakiire L, Kihembo C, Katushabe E, Natseri N, Nabukenya I, et al. Evaluation of integrated disease surveillance and response (IDSR) core and support functions after the revitalisation of IDSR in Uganda from 2012 to 2016. BMC Public Health. 2019 Jan 9;19:46. doi:10.1186/s12889-018-6336-2 PubMed PMID: 30626358; PubMed Central PMCID: PMC6327465.

28. World Health Organization. Measles vaccines: WHO position paper, April 2017 – Recommendations. Vaccine. 2019 Jan 7;37(2):219–22. doi:10.1016/j.vaccine.2017.07.066

29. Amodan BO, Ssendikwanawa E, Namayanja J, Opio BR, Biroma G, Morukileng J, et al. Epidemiological investigation of measles outbreak in a refugee settlement in Lamwo District, Uganda. Pan Afr Med J. 2025 Jul 25;51(Suppl 1):13. doi:10.11604/pamj.supp.2025.51.1.47672 PubMed PMID: 41211032; PubMed Central PMCID: PMC12595563.

30. Namulondo E, Ssemanda I, Komugisha M, Wako S, Nsubuga EJ, Kayiwa J, et al. Measles outbreak at a refugee settlement, Kiryandongo District, Uganda, July–October 2023. Vol. 9. 2024;9(1).

